# The structural covariance of reading-related brain regions in adults and children with typical reading skills and developmental dyslexia

**DOI:** 10.1101/2023.07.03.23292169

**Authors:** Amelie Haugg, Nada Frei, Christina Lutz, Sarah V. Di Pietro, Iliana I. Karipidis, Silvia Brem

## Abstract

Structural covariance (SC) is a promising approach for investigating brain organization within the domain of literacy and developmental disorders as it thought to reflect both functional and structural information. This study presents a first-of-its-kind exploration of SC in reading-related brain regions across different ages and reading abilities.

Whole-brain SC analyses were conducted for six key regions of the reading network, including an anterior and posterior subdivision of the visual word form area (VWFA). We compared SC matrices of typically reading adults (N=134) and children (N=110), and between typically reading children and children with dyslexia (N=68).

Our results showed significant associations between reading-related brain regions in typically reading adults. We observed significant SC between the posterior VWFA and the left occipital cortex, and between the anterior VWFA and the left superior temporal gyrus and left inferior frontal gyrus. Typical-reading adults and children did not differ significantly in SC. However, typically reading children demonstrated significantly higher SC between the inferior parietal lobule (IPL) and other reading-related brain regions than children with dyslexia.

Our findings provide support for a functional and structural division of the VWFA and underscore the crucial role of the IPL in fluent reading.

## Introduction

Reading is an essential skill in our society. It enables us to communicate and navigate, and it substantially influences our academic and professional development (Slavin, Lake, Chambers, Cheung, & Davis, 2009). Around 5-10% percent of the population, however, suffer from developmental dyslexia (DD) (Shaywitz, Shaywitz, Fletcher, & Escobar, 1990), a specific learning disorder characterized by significant impairments in reading performance which cannot be explained by deficits in intelligence or motivation (Shaywitz & Shaywitz, 2005).

DD has been associated with altered signals in various regions across the brain’s reading network (Maisog, Einbinder, Flowers, Turkeltaub, & Eden, 2008). During reading-related and phonological tasks, individuals with DD have been found to show significantly weaker brain activity within the visual word form area (VWFA) in the left ventral occipito-temporal cortex (vOTC) (Brem et al., 2020), the superior temporal gyrus (STG) (Ye, Rüsseler, Gerth, & Münte, 2017), and the left inferior parietal lobule (IPL) (Richlan, Kronbichler, & Wimmer, 2009). In contrast, other areas of the reading network were found to show increased activity in readers with DD as compared to typical readers. This includes hyperactivation in the precentral gyrus (PCG) which is associated with articulation (Brem et al., 2020; Cao et al., 2017) and the inferior frontal gyrus (IFG) which has been linked to higher-order reading processes such as memory and attention (Hoeft et al., 2007; Shaywitz et al., 1998; However, see contradicting results in Maisog et al., 2008). Hyperactivation in readers with DD has been suggested to be driven by compensatory mechanisms and increased recruitment of cognitive capacities during the reading process (Hancock, Richlan, & Hoeft, 2017).

Crucially, these key regions of the reading network have not only been associated with differences in functional brain activity but have also been found to show differences in brain structure when comparing individuals with DD to individuals with typical reading skills. In particular, individuals with DD demonstrate reduced grey matter volume in brain areas where they typically also show functional hypoactivation: A meta-analysis of studies investigating brain structure related to reading performance revealed a significant reduction of grey matter volume in the supramarginal gyrus/IPL, the STG, and the fusiform gyrus/VWFA (Linkersdörfer, Lonnemann, Lindberg, Hasselhorn, & Fiebach, 2012) when comparing poor readers to typical readers. In addition, the IFG has been found to show reduced grey matter volume in children with DD compared to typical-reading children (Jednoróg, Gawron, Marchewka, Heim, & Grabowska, 2014). In contrast, the PCG, which has been found to show functional hyperactivation in readers with DD, has been reported to demonstrate higher grey matter volume when comparing readers with DD to readers with typical reading skills (Jednoróg et al., 2015; Vinckenbosch, Robichon, & Eliez, 2005). However, it is important to note that the robustness of structural alterations in the brains of individuals with dyslexia has been challenged, mainly due to the fact that the majority of published results from older studies were derived from relatively small sample sizes making it even more important to clarify potential difference in larger samples (Jednoróg et al., 2015; Ramus, Altarelli, Jednoróg, Zhao, & Scotto di Covella, 2018).

While these findings provide useful insights into the functional and structural association of key brain regions with reading performance, they do not convey a deeper understanding of the complex interactions, dynamics, and connections between these regions. Consequently, new approaches are needed to investigate brain alterations in DD from a network perspective. In the last years, thus, much research has been performed to examine functional and effective connectivity within the brain’s reading network, shining light on the complex functional interplay of reading-related brain regions both in individuals with DD and typical reading skills (Di Pietro et al., 2023; Finn et al., 2014; Y. Li, Li, Yang, & Bi, 2022; Morken, Helland, Hugdahl, & Specht, 2017; Wang, Karipidis, Pleisch, Fraga-González, & Brem, 2020). These findings have also been complemented by studies using diffusion tensor imaging to investigate structural connections between key reading-related brain regions in children and adults with and without DD (Vandermosten, Boets, Wouters, & Ghesquière, 2012). However, to date, associations between grey matter volume in reading-related brain regions in general and, in particular, structural covariance, have not yet been investigated in the context of reading and DD.

Structural covariance is a measure of the associations of structural brain measures and describes how structural measures, such as grey matter volume, co-vary across the brain (Alexander-Bloch, Giedd, & Bullmore, 2013). As correlations between the sizes of different brain structures cannot be fully explained by brain size alone (Kennedy et al., 1998), it is assumed that structural covariance is driven by a meaningful brain organization principle. Indeed, studies suggest that structural covariance might be driven by both environmental and genetic factors (Mechelli, Friston, Frackowiak, & Price, 2005). Importantly, structural covariance presumably reflects structural as well as functional information (Alexander-Bloch, Giedd, et al., 2013). In specific, previous studies have shown that functional connectivity and white matter tracts are able to describe a large part of observed structural covariance in the brain (Alexander-Bloch, Raznahan, Bullmore, & Giedd, 2013; Gong, He, Chen, & Evans, 2012).

This strong coupling of functional activity levels and structural measures make structural covariance a particularly interesting and promising method to investigate brain organization in the context of literacy and developmental disorders. Indeed, a recent study found both increased and decreased structural covariance between brain regions involved in number processing in children with developmental dyscalculia, a specific learning disorder defined by impairments in arithmetic operations, as compared to typically developing children (Michels, Buechler, & Kucian, 2022).

Here, we report the first structural covariance findings on reading development. We aimed at answering the following questions: How is grey matter volume in core regions of the reading network associated with grey matter volume across the whole brain in the mature brain of typical readers? How does structural covariance change with brain maturation and increasing reading expertise from childhood to adulthood in typical readers? How do typical-reading children and children with dyslexia differ with regard to structural covariance of grey matter volume in these reading-related brain regions?

To address these questions we investigated the structural covariance of reading-related brain regions in adults and children with varying reading skills. First, we explored the structural covariance of key regions of the reading network in typical-reading adults to gain a better understanding of the potential interplay between these regions, but also between reading-related brain regions and the rest of the brain. To test for developmental effects, we then compared the structural covariance of reading-related brain regions between typical-reading adults and typical-reading children. Finally, we investigated differences in structural covariance between participants with typical reading skills and participants with DD.

Additionally, in accordance with previous studies suggesting that the VWFA encompasses two distinct subregions, the posterior VWFA which selectively responds to orthographic/perceptual aspects of text-related information and the anterior VWFA which is sensitive to lexical information (Caffarra, Karipidis, Yablonski, & Yeatman, 2021; Lerma-Usabiaga, Carreiras, & Paz-Alonso, 2018), we performed our analyses using two instead of one VWFA region of interest (ROI). This distinction allowed us to test whether the two VWFA subregions differed with respect to structural covariance in typical-reading adults, typical-reading children, and children with DD.

## Methods

### Participants

In total, data from 153 German-speaking, healthy adults (*M*=25.34, *SD* =4.08 years) and 201 German speaking, healthy children (*M*=9.84, *SD*=1.52 years) was collected for this study. All participants reported no psychiatric or neurological disorders with the exception of ADHD and dyscalculia and fulfilled the criteria for MR compatibility (e.g. no metal implants, no claustrophobia, no pacemakers, etc.). The children’s group was further divided into typical and poor readers based on their performance on a reading and decoding fluency test (Moll & Landerl, 2010). Children with a mean reading fluency score for words and pseudowords below or equal to the 16^th^ percentile were categorized as *children with DD* (N=68), children with a mean reading fluency score for words and pseudowords above or equal to the 25^th^ percentile were categorized as *typical-reading children* (N=110). For the adult group, only typical-reading participants with a mean reading fluency score for words and pseudowords above or equal the 25^th^ percentile were included in the final sample (N=134). A more detailed description of all participant groups is provided in Table 1. All data stem from studies approved by the Ethics Commission of the Canton of Zurich.

**Table 1:** Demographics and behavioral results of typical-reading children and adults, as well as children with dyslexia. MRI quality reflects a quality measure of the raw data as calculated by CAT12 for a scale from 1 to 100.

|  | Typical-reading adults | Typical-reading children | Children with DD | Comparison children with/without DD |
| --- | --- | --- | --- | --- |
| N | 134 | 110 | 68 |  |
| Handedness: l:r:b | 0:134:0 | 10:98:2 | 5:63:0 | $t(176)=1.11$<br>$p=0.27$ |
| Sex m:f | 49:85 | 52:58 | 32:36 | $t(176)=0.03$<br>$p=0.49$ |
| Nonverbal IQ (std) percentile | n.a. | 62.07 (17.22) | 56.08 (17.81) | $t(171)=2.28$<br>$p=0.02$ |
| Age in years (std)<br>Min – Max | 25.26 (4.01) | 9.61 (1.59)<br>6.88 – 12.09 | 10.21 (1.39)<br>7.47 – 12.16 | $t(176)=2.59$<br>$p=0.01$ |
| Reading fluency percentile (std) | 62.09 (21.55) | 59.35 (21.40) | 5.44 (4.40) | $t(176)=20.49$<br>$p<0.001$ |
| MRI quality percentage (std) | 87.06 (0.32) | 86.33 (1.20) | 86.34 (1.12) | $t(176)=0.05$<br>$p=0.96$ |

### Image acquisition

All structural scans were collected at the 3 Tesla Philips Achieva (Philips, Best, The Netherlands) MRI scanner of the Psychiatric University Hospital Zurich. The images were acquired with a 32-channel head coil and using the following scan parameters: repetition time (TR) = 6.8ms, echo time (TE) = 3.2ms, flip angle (FA) = 9°, field of view (FOV) 270 x 255 x 176 mm^3^, voxel size of 1 x 1 x 1 mm^3^, 176 slices acquired in sagittal slice orientation. A detailed description of all sequence information can be found on our Open Science Framework repository (see exam cards folder at: https://osf.io/5bekd/).

### MRI preprocessing and quality control

MRI preprocessing was performed using MATLAB2020b (www.mathworks.com), SPM12 (https://www.fil.ion.ucl.ac.uk/spm/software/spm12/), and the CAT12 toolbox (https://neuro-jena.github.io/cat/). Following CAT12’s default segmentation pipeline, preprocessing steps included internal interpolation, denoising, affine preprocessing, local adaptive segmentation, adaptive maximum a posterior segmentation, partial volume segmentation, skull-stripping, spatial normalization, and white matter hyperintensity correction. To account for anatomical differences in children as compared to adults, tissue probability maps (TPM) for the two pediatric datasets were custom-made for the respective age group using the Template-O-Matic Toolbox (TOM) (https://neuro-jena.github.io/software.html#tom). The two age-adjusted TPMs can be downloaded from our Open Science Framework repository (https://osf.io/5bekd/). After preprocessing, CAT12’s quality assessment measures were used to identify participants with anatomical images of poor quality (quality ratings below 75%) which were excluded from further analyses. On average, anatomical images received data quality ratings of 87.06% for adults and 86.38% for children. Finally, we performed smoothing with an 8mm kernel. The scripts used for data preprocessing can be found in this study’s Open Science Framework repository (https://osf.io/5bekd/).

### Regions of interest

Regions of interest (ROIs) for structural covariance analyses were created as spheres with a radius of 4mm using Marsbar (https://marsbar-toolbox.github.io/). Spheres were built around left-hemispheric coordinates taken from the literature (Di Pietro et al., 2023; Lerma-Usabiaga et al., 2018) and, in the case of the STG based on Neurosynth meta-analysis (neurosynth.org, search term “superior temporal”). As recent research suggests that the VWFA can be divided into two separate subregions linked to different text-processing functions, we created two separate VWFA ROIs: The anterior VWFA_lex (-42, -58, -10), which has been associated with more lexical functions, and the posterior VWFA_per (-39, -72, -8), which has been associated with more perceptual functions (Lerma-Usabiaga et al., 2018). The other ROIs encompassed the STG (-52, -22, 6), PCG (-44, 6, 30), IFG (-56, 12, 15), and IPL (-40, -48, 42). All coordinates are provided in MNI space and the spherical ROIs used in our analyses can be downloaded from this study’s repository on the Open Science Framework (https://osf.io/5bekd/).

### Seed-based analysis to explore structural covariance in the reading network

First, we performed a seed-based analysis to investigate which voxels across the whole brain were significantly associated with grey matter volume in the five seed ROIs in typical-reading adults. To perform the seed-based analyses, we extracted the mean grey matter volume from each ROI sphere (see above for details regarding ROIs). Then, we performed a second-level general linear model (GLM) using the participants’ total intracranial volume (TIV) as a covariate of no interest and the ROI’s mean extracted grey matter volume as a covariate of interest, similar to Li and colleagues (X. Li et al., 2013). This analysis was performed separately for each of the five seed ROIs. The output of these analyses was interpreted using an initial threshold of p<0.001 and a cluster threshold of p<0.05 (family-wise error-corrected).

### Structural covariance analysis to investigate structural development

To investigate structural covariance between reading-related brain regions in typical-reading children and adults, we created one covariance matrix for each group using MATLAB2022a. Each structural covariance matrix was based on mean grey matter volume of the reading-related brain regions that were used as seeds in the GLM analyses of adults and that demonstrated significant associations with other brain regions in those GLM analyses (i.e., VWFA_lex, VWFA_per, STG, IFG, IPL). All mean grey matter volume values were corrected for TIV before the calculation of the covariance matrices. In order to calculate whether mean structural covariance matrices differed significantly between the two groups, we performed permutation tests with 1000 permutations. When two matrices differed significantly, we additionally performed permutation tests as post-hoc tests for all pairs of ROIs. Finally, for completeness, we also performed structural covariance analyses across the whole brain for 140 ROIs taken from the Neuromorphometrics parcellations (Neuromorphometrics, Inc., Somerville, MA). Again, we calculated structural covariance matrices for each group and compared them using permutation tests with 1000 permutations.

### Structural covariance analysis to investigate structural alterations with reading skills

Finally, we investigated structural alterations associated with reading skills in children with and without reading impairments. To do so, structural covariance analysis was performed using the same analysis steps as for the comparison between typical-reading adults and children. In addition, we corrected for age and non-verbal intelligence.

## Results

### Whole-brain seed-based analysis of typical-reading adults

We performed GLMs to investigate associations in grey matter volume between key regions of the reading network and voxels in the rest of the brain. Tables with coordinates of all significant associations resulting from the GLMs can be found in the Supplemental Material (Tables S1-S6).

In typical-reading adults, we observed significant associations (FWE-corrected with a p-value below 0.05) between the left-hemispheric VWFA_lex seed and voxels in the right vOTC, as well as in the left STG, and left IFG (see Figure 1, Table S1 in Supplemental Material). For the VWFA_per seed, we observed significant associations with the left ventral occipital cortex (vOCC), posterior parts of the left vOTC, and the left middle occipital gyrus (see Figure 1, Table S2 in Supplemental Material).

**Figure 1:**
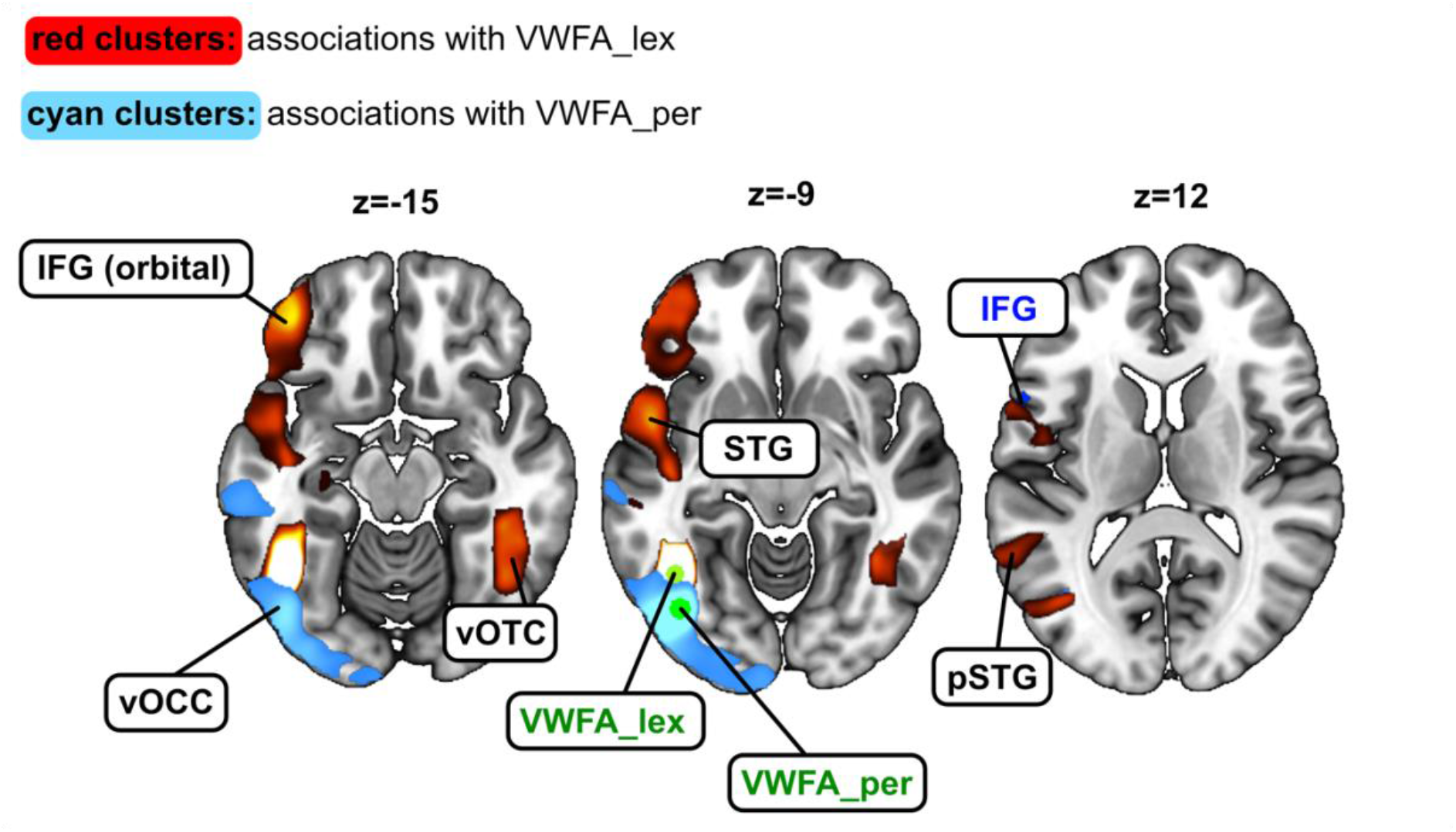
Structural covariance maps of grey matter volume in the lexical part of the visual word form area (VWFA_lex) and the perceptual part of the visual word form area (VWFA_per). For the VWFA_lex seed, we observed significant structural covariance with the right ventral occipito-temporal cortex (vOTC), the left inferior frontal gyrus (IFG), and the anterior part as well as the posterior part of the superior temporal gyrus (STG) (see red clusters). For the VWFA_per seed, we observed significant associations with the left ventral occipital cortex (vOCC) (see cyan clusters). The seed regions (here: VWFA_lex and VWFA_per) are depicted in green, and relevant seed regions of the other analyses in this study are depicted in blue.

For the left STG seed, adult readers showed significant associations bilaterally with large parts of the STG and with the right superior frontal gyrus (SFG) (see Figure 2A, Table S3 in Supplemental Material). When using the left IPL as a seed, we observed significant grey matter volume associations with the right IPL, the left IFG, the left orbitofrontal cortex (OFC) as well as parts of the prefrontal cortex (PFC), the posterior cingulate cortex (PCC), and bilaterally in the caudate (see Figure 2B, Table S4 in Supplemental Material). For the seed in the left IFG, we observed significant structural covariance between the grey matter volume in the seed and a connected cluster containing both the PFC and left PCG, and between the IFG seed and the PCC (see Figure 2C, Table S5 in Supplemental Material). Finally, for the left PCG seed, we did not find any significant associations of grey matter volume with clusters that were not part of the left PCG (see Table S6 in Supplemental Material).

**Figure 2:**
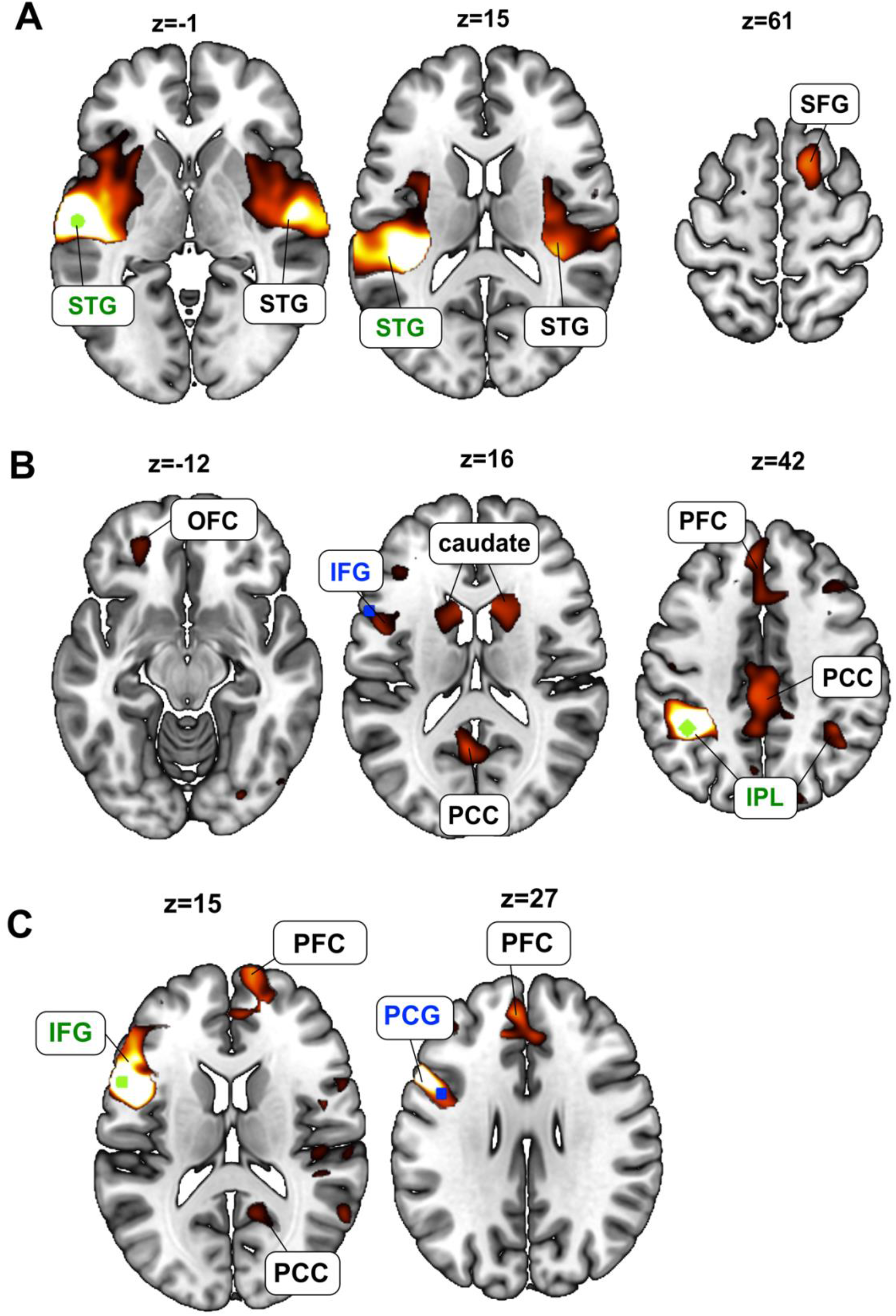
Structural covariance map of grey matter volume in the left superior temporal gyrus, left inferior parietal lobule, and left inferior frontal gyrus. A. We observed significant associations between grey matter volume in the left and right superior temporal gyrus (STG), as well as between the left STG and the right superior frontal gyrus (SFG). The seed region (here: STG) is depicted in green. **B.** We observed significant associations between grey matter volume in the left and right inferior parietal lobule (IPL), as well as between the left IPL and left inferior frontal gyrus (IFG), orbitofrontal cortex (OFC), bilateral caudate, prefrontal cortex (PFC), and posterior cingulate cortex (PCC). The seed region (here: IPL) is depicted in green, relevant seed regions of other analyses in this study are depicted in blue. **C**. We observed significant associations between grey matter volume in the left inferior frontal gyrus (IFG) and the left precentral gyrus (PCG), the prefrontal cortex (PFC), and posterior cingulate cortex (PCC). The seed region (here: IFG) is depicted in green, relevant seed regions of other analyses in this study are depicted in blue.

### Structural covariance in typical-reading children and adults

In the next analysis, we investigated the typical development of structural covariance between reading-related brain regions by comparing structural covariance matrices between typical-reading children and typical-reading adults. As the PCG did not show any significant associations to other brain regions in the seed-based whole brain analysis of adult participants, we decided to exclude the PCG from the focus of this structural covariance analysis. We did not find a significant difference in mean structural covariance of reading-related brain regions between typical-reading children and adults (p=0.55) (see Figure 3 and Supplemental Material Figure S1). For completeness, we repeated this analysis for 140 ROIs across the whole brain (see Supplemental Material Figure S2 and S3) and did not find a significant difference in overall structural covariance of the whole brain either (p=0.13).

**Figure 3:**
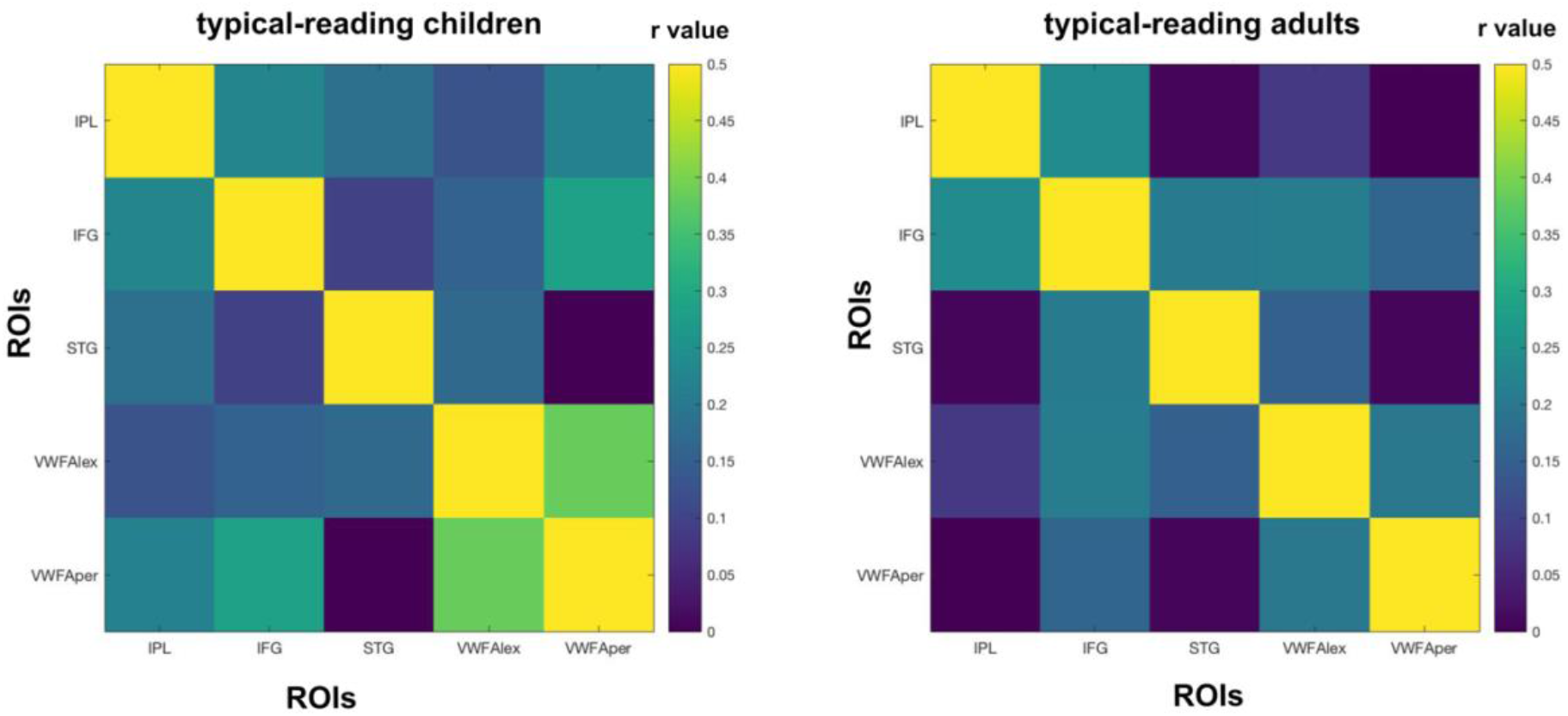
Structural covariance of key regions of the reading network in typical-reading children and adults. We did not find a significant difference in the mean structural covariance of reading-related brain regions between typical-reading children and adults. The color of each cell reflects the structural covariance of the corresponding column and row ROI. A visual representation of the difference between the two matrices can be found in the Supplemental Material (Figure S1). Abbreviations: Inferior Parietal Lobule (IPL), Inferior Frontal Gyrus (IFG), Superior Temporal Gyrus (STG), lexical Visual Word Form Area (VWFA_lex), perceptual Visual Word Form Area (VWFA_per).

### Structural covariance in typical-reading children and children with dyslexia

Then, we compared the structural covariance of reading-related brain regions between typical-reading children and children with DD. We observed a significant difference in overall structural covariance of reading-related brain regions (p=0.015) which suggest an overall higher structural covariance among regions of the reading network in typical-reading children. To further investigate whether this difference was driven by the covariance between specific brain regions, we performed pairwise comparisons. These comparisons revealed significantly higher structural covariance for typical-reading children compared to children with DD between IPL-STG (p=0.005), IPL-VWFA_per (p=0.007), and a trend for IFG-VWFA_per (p=0.09). No significant differences were found for the IPL-IFG (p=0.11), IPL-VWFA_lex (p=0.15), IFG-STG (p=0.94), IFG-VWFA_lex (p=0.32), STG-VWFA_lex (p=0.89), STG-VWFA_per (p=0.64), and the VWFA_lex-VWFA_per (p=0.84). Only the IPL-STG and the IPL-VWFA_per structural covariance comparisons survived a multiple comparison correction using Benjamini & Hochberg false discovery rate (FDR) correction. Figure 4 provides an overview of the structural covariance of key reading-related brain regions in both typical-reading children and children with dyslexia (see Figure 5 for a visualization of the differences).

**Figure 4:**
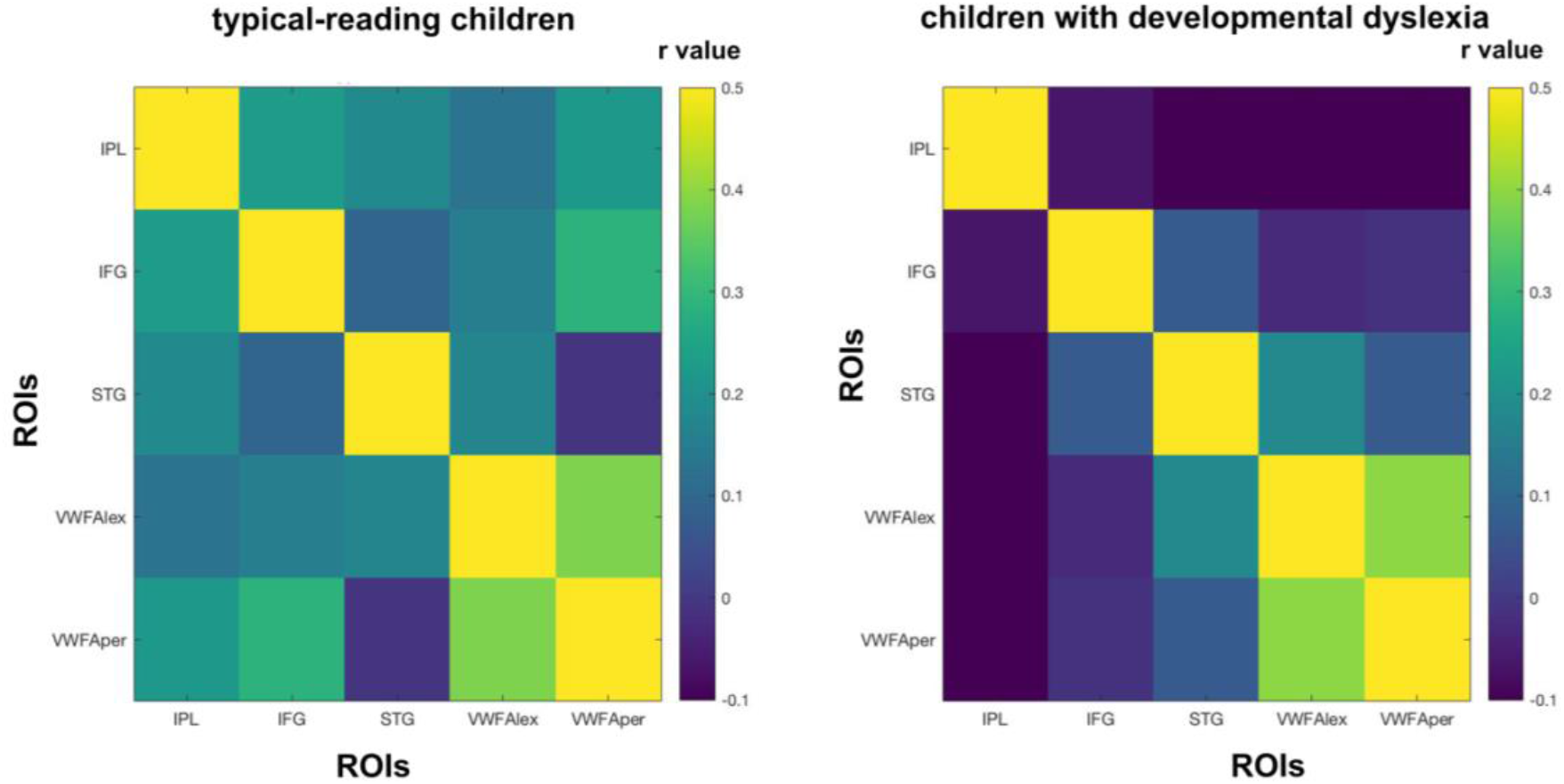
Structural covariance of key regions of the reading network in typical-reading children and children with dyslexia. On average, typical-reading children showed higher structural covariance between reading-related brain regions than children with dyslexia (p=0.015). This was mainly driven by higher levels of structural covariance of the inferior parietal lobule (IPL) with other regions of the reading-network, such as between IPL-STG and between IPL-VWFA_per. The color of each cell reflects the structural covariance of the corresponding column and row ROI. Abbreviations: inferior parietal lobule (IPL), inferior frontal gyrus (IFG), superior temporal gyrus (STG), lexical visual word form area (VWFA_lex), perceptual visual word form area (VWFA_per).

**Figure 5:**
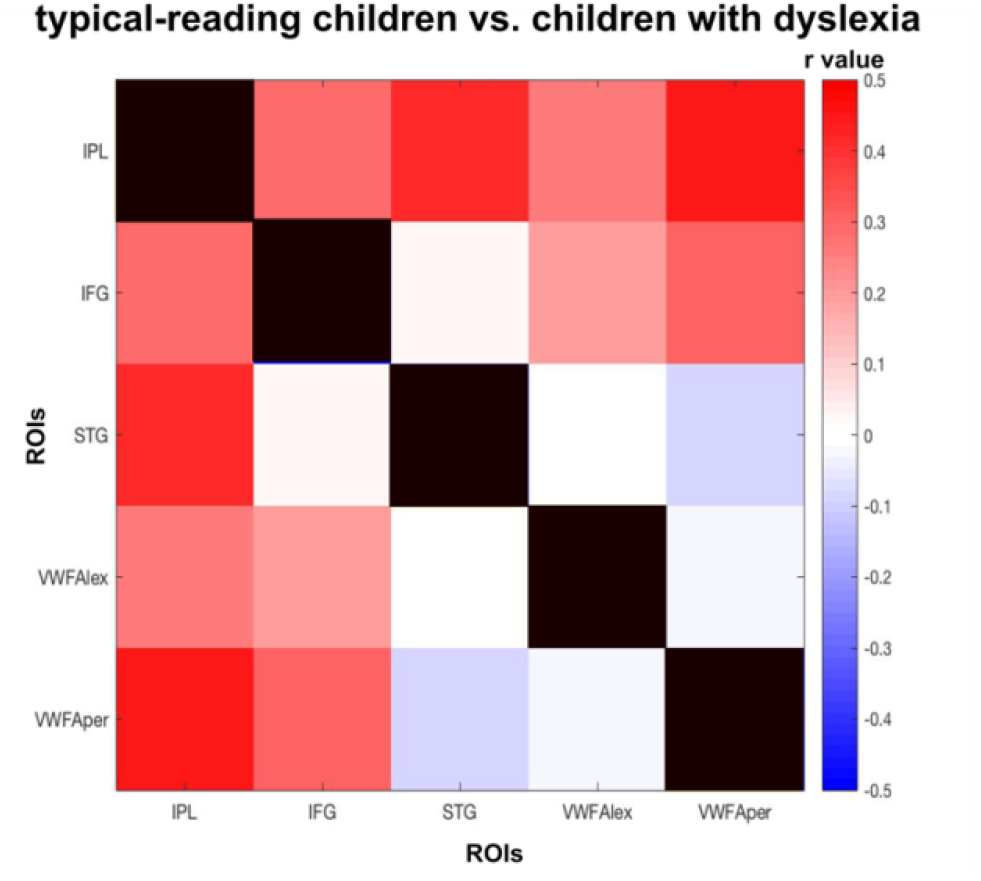
Difference in structural covariance of reading-related brain regions between typical-reading children and children with dyslexia. We found a significant difference between typical-reading children and children with dyslexia for structural covariance of reading-related brain regions (p=0.015). The color of each cell reflects the difference in structural covariance (typical-reading children minus children with dyslexia) of the corresponding column and row ROI.

We did not find a significant difference in mean structural covariance between typical-reading children and children with dyslexia when performing structural covariance analyses using 140 ROIs from a whole-brain parcellation (p=0.35) (see Figures S4 and S5 in the Supplemental Material), suggesting that the difference in structural covariance may be mainly restricted to areas relevant for reading..

Finally, we investigated whether typical-reading children as well as children with dyslexia would demonstrate a differentiation between VWFA_per and VWFA_lex with regard to associations to other brain regions on a whole brain level. For children with dyslexia, both the VWFA_lex and the VWFA_per only showed significant associations with the vOTC, either unilateral (VWFA_per) or bilateral (VWFA_lex) (see Figure 6 and Supplemental Material Tables S7 and S8). For typical-reading children, we found significant associations of the VWFA_lex with the posterior left STG and the right insula, and of the VWFA_per with the right IPL (see Figure 7 and Supplemental Material Tables S9 and S10).

**Figure 6:**
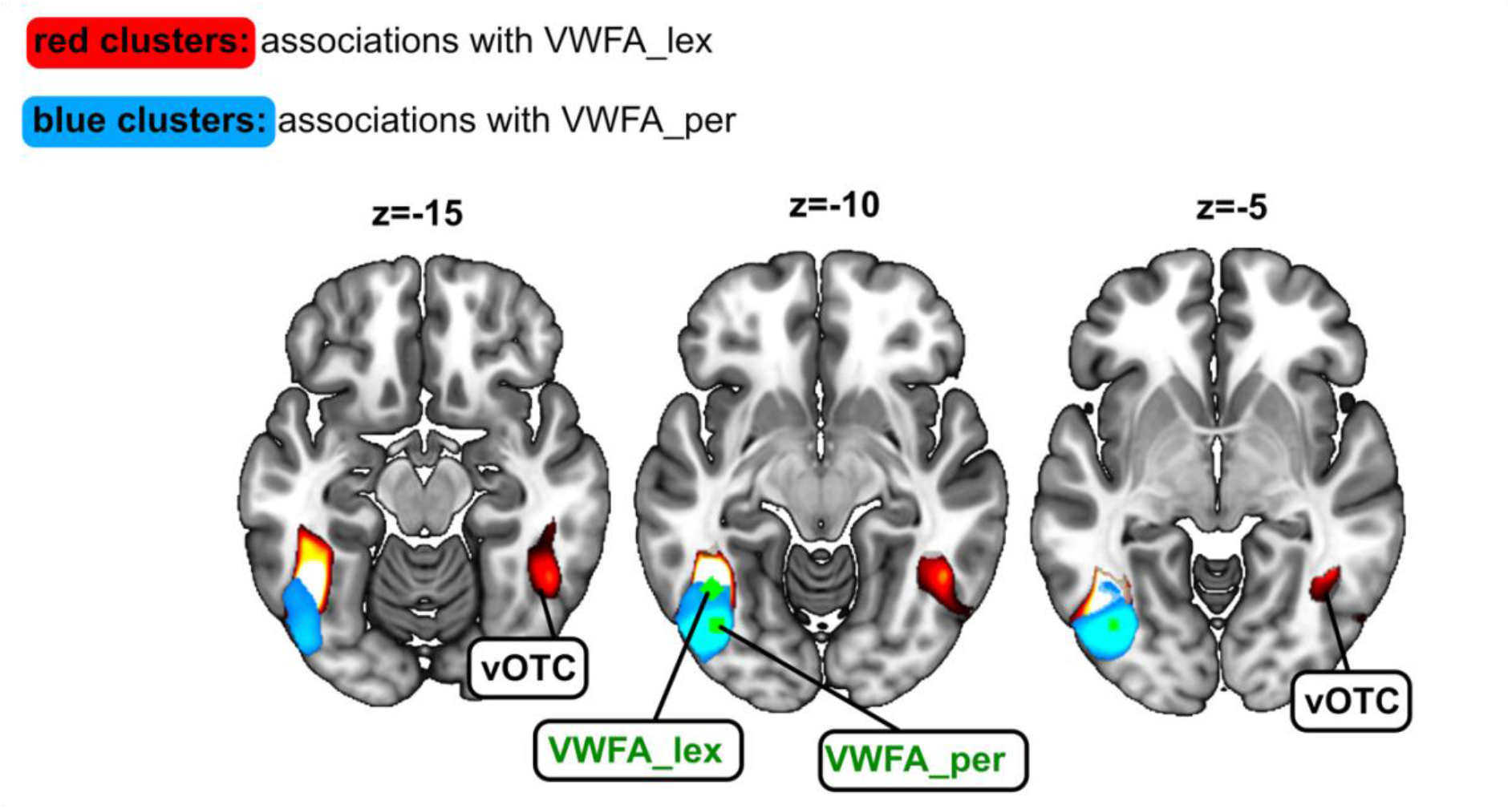
Structural covariance map of grey matter volume in the lexical part of the visual word form area (VWFA_lex) and the perceptual part of the visual word form area (VWFA_per) in children with dyslexia. For the VWFA_lex seed, we observed significant associations with the right ventral occipitotemporal cortex (vOTC). For the VWFA_per seed, we did not observe any significant associations with brain regions other than the seed area.

**Figure 7:**
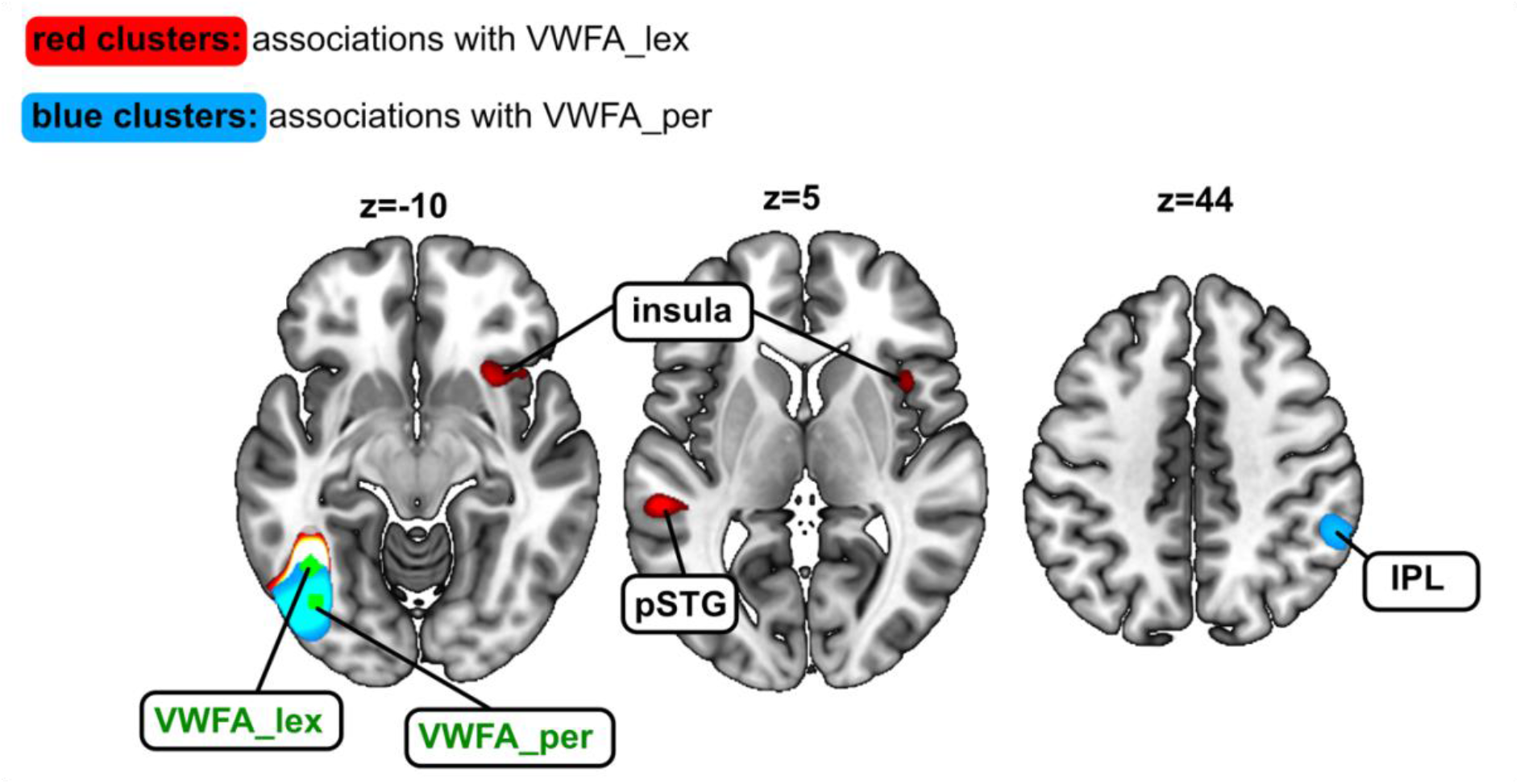
Structural covariance map of grey matter volume in the lexical part of the visual word form area (VWFA_lex) and the perceptual part of the visual word form area (VWFA_per) in typical-reading children. For the VWFA_lex seed, we observed significant associations with the right insula as well as the posterior part of the left superior temporal gyrus (pSTG). For the VWFA_per seed, we observed significant associations with the right inferior parietal lobule (IPL). The seed regions (here: VWFA_lex and VWFA_per) are depicted in green.

## Discussion

In the present work, we investigated the association of grey matter volume between the key regions of the brain’s reading network and the rest of the brain by performing structural covariance analyses with a series of GLMs, each with one key region’s grey matter volume added as a covariate. This focus on key regions of the reading network distinguishes our analyses from previous studies investigating structural covariance and structural associations without any focus on reading processes. In our analyses, we observed significant associations between reading-related brain regions as well as between reading-related brain regions and regions not classically related to the reading process (such as the bilateral caudate).

### Structural covariance of VWFA subregions in typical-reading adults

In a first analysis, we explored the structural covariance of two different VWFA seeds, namely one seed in the anterior part of the VWFA (VWFA_lex) and one seed in the posterior part of the VWFA (VWFA_per). Text recognition in the fusiform gyrus is hierarchically organized with increased sensitivity to orthographic properties of written text in more anterior than posterior parts (Vinckier 2007, Brem 2006, for a review: Caffarra et al. 2021). Recent studies suggest that, rather than being a single region of the brain’s reading network, the VWFA is divided into two functionally distinct anterior and posterior subregions associated with lexical and perceptual processing, respectively. In particular, Lerma-Usabiaga and colleagues provided functional, structural, and behavioral evidence for this distinction (Lerma-Usabiaga et al., 2018). However, thus far, it has not been investigated whether this distinction is also found for structural covariance. Here, in line with Lerma-Usabiaga and colleagues, we found significant structural covariance of the perceptual part of the VWFA only with visual brain regions in the occipital cortex. In contrast, for the lexical part of the VWFA, we observed structural associations with large clusters of key regions associated with reading, such as the STG and the IFG. Consequently, our findings provide further support for the subdivision of the VWFA into a lexical and a perceptual part. In addition, our findings are in line with studies investigating functional connectivity of the two VWFA subregions (van der Mark et al., 2011; Yablonski, Karipidis, Kubota, & Yeatman, 2023). In a seed-based whole-brain analysis at an individual level, Yablonski and colleagues found strong functional correlations between the perceptual VWFA seed and visual regions. In contrast, when using the lexical VWFA as a seed for the functional connectivity analysis, they observed stronger functional connectivity to the inferior frontal cortex (Yablonski et al., 2023).

As the VWFA has been found to be highly specialized in reading processes in literate individuals (Brem et al., 2020; Cohen et al., 2000) and as our results closely reflect previous findings of the VWFA_lex and VWFA_per, it is very likely that the structural associations we observed between the two VWFA seeds and other regions of the brain are indeed driven by reading processes rather than other brain functions. It can be expected that the association of the VWFA_per with the visual cortex is linked to perceptual processing during reading, involving both VWFA_per and other visual regions. Similarly, it is likely that associations of the VWFA_lex with other brain regions are linked to the processing of lexical stimuli. Interestingly, we also observed a significant association between VWFA_lex and the left STG, a connection which has not been observed by Lerma-Usabiaga et al. and Yablonski et al (Lerma-Usabiaga et al., 2018; Yablonski et al., 2023). This could be related to the language the study was conducted in. Our participants were native German speakers, while Lerma-Usabiaga and Yablonski investigated data of English speakers. As the STG is an important brain region for phonological processing and audio-visual integration during reading processes (Karipidis, Pleisch, Di Pietro, Fraga-González, & Brem, 2021) and demonstrates hypoactivation in readers with DD (Blau, van Atteveldt, Ekkebus, Goebel, & Blomert, 2009; Ye et al., 2017), it is, again, likely that this structural covariance finding can be linked to reading processes. Indeed, other studies have shown that functional connectivity between the VWFA and the STG is correlated with reading performance, such that better reading skills coincide with stronger functional connectivity (Chai et al., 2016).

### Structural covariance of reading-related brain regions in typical-reading adults

In addition to the two VWFA subregions, we also explored associations with other key regions related to reading. However, for several of these ROIs, structural covariance appeared to be less dominantly linked to reading and lexical processing. For instance, when specifically investigating structural associations of the STG as a seed, we did not find any significant associations with typical key regions of the reading network, but only significant associations with the STG in the right hemisphere and the SFG. As the STG, in contrast to the VWFA, is involved in a rather wide range of auditory functions and multisensory processing not exclusively related to reading, it would not be surprising if a more general auditory processing was driving structural associations of the STG (Chang et al., 2018; Yi, Leonard, & Chang, 2019).

Indeed, this reasoning of additional functions other than reading might also explain our findings for the IPL seed. As the IPL is part of the default mode network, a robust and strongly-connected network of brain regions involved in mind wandering that is consistently found in functional connectivity analyses during the resting state (Buckner, Andrews-Hanna, & Schacter, 2008; Raichle & Snyder, 2007), structural associations to other regions of the default mode network, such as the PFC and the PCC are not surprising. Nevertheless, we also observed a significant association between the left IPL seed and the left IFG. Given that this finding was clearly left-lateralized, this finding might be driven by reading- or language-related processes. Similarly to the reading-related structural associations discussed above, previous studies have found a significant association between increased functional connectivity between the IPL and the IFG and gains in phonological processing skills in children (Yu et al., 2018). Further, a recent effective connectivity study showed increased feedback-connectivity from IPL to the VWFA in children with dyslexia, suggesting a role of the IPL in guiding attention to letters, for example during decoding processes (Di Pietro et al., 2023).

Similarly as for the IPL, we also observed significant associations with the PCC and the PFC for the IFG seed. Here, the association with the PFC needs to be interpreted with caution as the PFC was part of the cluster that also contained the IFG seed and might, therefore, be driven solely by spatial proximity rather than a meaningful underlying organization based on brain function. This also applies to the significant association with the PCG which, too, was located in the same cluster as the seed sphere.

Finally, the PCG seed only demonstrated significant associations with the cluster that also contained the seed sphere. Again, this association was most likely only driven by spatial proximity rather than similar functions and, consequently, in our sample, the PCG appeared to be isolated from other brain regions with regard to structural covariance. However, it is possible that associations to other brain regions might be found for larger sample sizes.

### Structural covariance of reading-related brain regions in typical-reading children and adults

To investigate the comparability between typical-reading children and adults, we performed permutation tests to assess potentially significant differences in structural covariance of reading-related brain regions between the two groups. Here, we did not observe any significant differences, indicating that children who have learned how to read and possess typical reading skills already exhibit the basic structural associations related to reading, similar to adult readers. Interestingly, the two groups did not differ even after when repeating this analysis for 140 ROIs across the whole brain. Our results are not in line with previous studies that observed differences in structural covariance related to development (DuPre & Spreng, 2017; X. Li et al., 2013; Zielinski, Gennatas, Zhou, & Seeley, 2010). For instance, Zielinski and colleagues found changes in structural covariance across different ages from childhood to adolescence (Zielinski et al., 2010). However, they did not correct for TIV, which could be a confounding factor in structural covariance analyses across ages. In addition, they did not perform statistical testing to compare structural covariance values between the groups, but rather compared the number of voxels associated with each seed, so comparability to our results is limited. Next, we should also note that our groups of children included children from ages 6.88 – 12.16 years, we thus pooled children’s data of a rather broad age range. One could hypothesize that when comparing children within a narrower age range, such as younger children versus older children or younger children versus adults, more distinct differences would likely emerge due to developmental effects. Also, it should be noted that most structural covariance analyses investigating development in children and adults focus on specific ROIs or networks, while our analysis was performed for 140 ROIs covering the whole brain thus providing a more global measure. Consequently, our results do not exclude the possibility that specific ROIs might, indeed, demonstrate significant developmental effects.

### Structural covariance analyses of reading-related brain regions in typical-reading children and children with DD

One of the main aims of this study was to investigate and compare structural covariance in typical-reading children and children with dyslexia. The two groups did not differ significantly in overall structural covariance when we used 140 ROIs across the whole brain to calculate the structural covariance matrices. This indicates that it is unlikely that reading impairments change associations of grey matter volume on a bigger scale across the whole brain. However, when we focused our analysis on reading-related brain regions, we found a significant difference between typical-reading children and children with dyslexia. Specifically, we observed significantly stronger structural covariance between the IPL and other reading-related brain regions in typical-reading children as compared to children with DD. This finding provides further evidence that structural covariance between reading-related brain regions, to some extent, reflects reading performance and functional reading processes in the brain. Consequently, future studies investigating potential structural biomarkers of developmental dyslexia and reading impairments should not only focus their analyses on brain structures per se but, also, consider interactions between these structures in typical and patient populations.

Further, we investigated whether specific brain structures were driving these differences between typical-reading children and children with dyslexia. We found that, in particular, the left IPL appeared to show stronger associations with other regions in typical-reading children as compared to children with DD. The left IPL, containing the angular gyrus and the supramarginal gyrus, has been found to play an important role in context-dependent integration during reading (Branzi, Pobric, Jung, & Ralph, 2021) as well as phonological processing (Sliwinska, Khadilkar, Campbell-Ratcliffe, Quevenco, & Devlin, 2012), and general word processing (Stoeckel, Gough, Watkins, & Devlin, 2009). Thus, the association of grey matter volume in the IPL with grey matter volume in these other reading-related brain regions is likely generated by the interplay of these regions during the processing of words and lexical stimuli.

This idea is further supported by the fact that the left IPL also demonstrates reading-related functional connectivity to other brain regions. For instance, an early study by Horwitz and colleagues using PET imaging discovered an absence of functional connections between the left IPL and other reading-related brain regions in readers with DD as compared to typical readers. Similarly to our results, they found weaker association of the left IPL with the left IFG, the VWFA, and the left STG (Horwitz, Rumsey, & Donohue, 1998). These findings are also in line with recent studies using fMRI which observed alterations in functional and effective connectivity between the left IPL and other reading-related brain regions. For instance, van der Mark and colleagues observed deficient functional connectivity between the left IPL and the VWFA in children with dyslexia as compared to typical-reading children (van der Mark et al., 2011). Comparable findings have been observed for effective connectivity in a recent study by Di Pietro et al. who observed differences in effective connectivity between the IPL and the VWFA when comparing children with typical reading skills and with dyslexia. Independent of the age of the children, they observed a diminished inhibitory connection from the IPL to the VWFA in readers with DD (Di Pietro et al., 2023). Taken together, these results emphasize the importance of the left IPL for lexical processing and the strong interplay between the left IPL and other regions of the reading network.

### Structural covariance of VWFA subregions in typical-reading children and children with dyslexia

Finally, we performed an analysis to investigate whether VWFA_lex and VWFA_per would demonstrate similar associations in children as we found for typical-reading adults. For children with dyslexia, we observed a significant association of the VWFA_lex with the right vOTC, but no significant associations with any brain regions other than the seed region for the VWFA_per. Based on these findings, it can be assumed that the differentiation between the anterior and posterior subregion within the VWFA is not yet present in our sample of children with DD with regard to structural covariance measures. We observed a unilateral association for the VWFA_per and a bilateral association for the VWFA_lex similarly to our findings for typical-reading adults. This observation of unilateral or bilateral structural covariance is likely unrelated to reading processes and might reflect the general architecture of the primary and secondary visual cortex.

For typical-reading children, the VWFA_per seed did not show any significant associations to the right vOTC, but we observed an association to the right IPL. Associations of the VWFA_per with inferior parietal regions are in line with previous findings on functional connectivity. For instance, Yablonski and colleagues reported significant functional connectivity patterns between the VWFA_per and visual as well as inferior parietal regions in children (Yablonski et al., 2023). For the VWFA_lex seed, similarly to the results of typical-reading adults, we found significant associations to the posterior STG. However, we did not find any significant structural association to the IFG, but rather to the right insula. It is less likely that this association to the insula is linked to primary reading processes (Uddin, Nomi, Hebert-Seropian, Ghaziri, & Boucher, 2017). However, as our samples of children contain fewer data points than the adult sample, differences between the groups might also reflect, to some extent, differences in power due to smaller sample sizes. Consequently, further studies will be needed to investigate this research question in a larger sample and particularly in more narrowly defined age ranges of typical-reading children and children with dyslexia. Overall, our findings for structural covariance of the two VWFA subregions in children demonstrate differences between the two ROIs for typical-reading children, similar to the findings of Yablonski and colleagues, but not for children with dyslexia. Future investigations will be needed to investigate whether a differentiation of structural covariance of the two VWFA subregions is present in adults with dyslexia or whether the lack of such a differentiation might be a general deficit in individuals with dyslexia.

### Limitations

We used 4mm spheres as seed regions for our whole-brain analyses, similar to previous structural covariance analyses (X. Li et al., 2013). On the one hand, this approach made our analyses more comparable to this previous study, on the other hand, however, it made it more likely that a single sphere might not capture the targeted brain region optimally due to its small size and the interindividual variability in brain anatomy. Individual definition of seeds in target areas using a functional localizer may help to overcome this challenge, but data from a functional localizer was not available for the current analysis.

## Conclusion

Our findings show that key regions of the reading network demonstrated significant structural associations in typical-reading adults. While these associations between reading-related brain regions did not differ significantly between typical-reading children and adults, we found significant differences between typical-reading children and children with dyslexia. In particular, associations between the left IPL and other reading-related brain regions were significantly stronger for children with typical reading skills than for children with dyslexia, emphasizing the importance of the left IPL and its interplay with other brain regions for lexical processing and reading.

We also compared the structural covariance between the lexical and perceptual VWFA in adults and children. Our findings support the notion that these two subregions are associated with different functions as well structural connections. In adults we found a strong structural covariance between the lexical VWFA and the left IFG, which was not yet evident in the children’s sample, suggesting that the structural and functional development of the brain’s reading circuit requires many years to mature.

Overall, our results provide evidence that reading performance and reading difficulties are likely associated with differences in the structural covariance of key reading brain regions. This highlights the importance of not only investigating structural measures of specific reading-related brain regions but also the associations between structural measures among these regions, when investigating brain structures linked to impaired and typical reading in the brain.

## Data Availability

All data produced in the present study are available upon reasonable request to the authors

## Acknowledgements

This study was funded by NCCR Evolving Language with the Swiss National science Foundation Agreement #51NF40-180888, and the Fonds für wissenschaftliche Zwecke im Interesse der Heilung von psychischen Krankheiten. The authors declare no conflicts of interest.

