## Supplemental Material for "The structural covariance of reading-related brain regions in adults and children with typical reading skills and developmental dyslexia"

*Whole-brain seed-based analyses of typical-reading adults*

| cluster | | peak | Coordinates (MNI) | | | anatomy |
|---|---|---|---|---|---|---|
| **p(FWE-corr)** | **equivk** | **T** | **x** | **y** | **z** | |
| <0.001 | 4888 | 86.96 | -44 | -58 | -10 | seed (VWFA_lex) in left vOTC |
| | | 5.35 | -58 | -56 | -26 | |
| | | 4.72 | -40 | -69 | 10 | |
| <0.001 | 7744 | 5.98 | -42 | 44 | -14 | left IFG and left STG |
| | | 5.5 | -54 | 8 | -6 | |
| | | 5.46 | -51 | 2 | -10 | |
| 0.001 | 2211 | 5.55 | 48 | -46 | -21 | right vOTC |
| | | 5.14 | 42 | -39 | -15 | |
| | | 5.1 | 39 | -51 | -12 | |

**Table S1: Brain regions with grey matter volume associated with grey matter volume in the lexical Visual Word Form Area (VWFA_lex).** Initial threshold p<0.001. Abbreviations: ventral occipito-temporal cortex (vOTC), Inferior Frontal Gyrus (IFG), Superior Temporal Gyrus (STG).

| cluster | | peak | Coordinates (MNI) | | | anatomy |
|---|---|---|---|---|---|---|
| **p(FWE-corr)** | **equivk** | **T** | **x** | **y** | **z** | |
| <0.001 | 4355 | 75.83 | -42 | -72 | -8 | seed (VWFA_per) including left visual cortex |
| | | 5.65 | -36 | -88 | -14 | |
| | | 4.48 | -6 | -99 | -4 | |
| 0.028 | 1081 | 4.34 | -51 | -76 | 24 | left middle occipital gyrus |
| | | 4.32 | -30 | -82 | 42 | |
| | | 3.54 | -36 | -66 | 28 | |

**Table S2: Brain regions with grey matter volume associated with grey matter volume in the perceptual Visual Word Form Area (VWFA_per).** Initial threshold p<0.001. Abbreviations: Visual Word Form Area (VWFA)

| cluster | | peak | Coordinates (MNI) | | | anatomy |
|---|---|---|---|---|---|---|
| **p(FWE-corr)** | **equivk** | **T** | **x** | **y** | **z** | |
| <0.001 | 17860 | 94.03 | -58 | -16 | 0 | seed (left STG) |
| | | 6.26 | -57 | -36 | 18 | |
| | | 5.02 | -33 | -4 | 8 | |
| <0.001 | 15146 | 8.6 | 60 | -18 | 6 | right STG |
| | | 8.34 | 56 | -14 | 0 | |
| | | 7.32 | 46 | -20 | 4 | |
| 0.025 | 1106 | 5.43 | 15 | 14 | 64 | SMA |

**Table S3: Brain regions with grey matter volume associated with grey matter volume in the left Superior Temporal Gyrus (STG).** Initial threshold p<0.001. Abbreviations: Supplementary Motor Area (SMA).

| cluster | | peak | Coordinates (MNI) | | | anatomy |
|---|---|---|---|---|---|---|
| **p(FWE-corr)** | **equivk** | **T** | **x** | **y** | **z** | |
| <0.001 | 3185 | 61.86 | -40 | -48 | 44 | seed (left IPL) |
| 0.02 | 1184 | 5.43 | 42 | 21 | 34 | right IPL |
| | | 4.83 | 51 | 15 | 30 | |
| | | 3.48 | 34 | 22 | 44 | |
| 0.003 | 1798 | 5.24 | -36 | 21 | 32 | PFC, including left IFG |
| | | 4.75 | -33 | 32 | 22 | |
| | | 4.41 | -48 | 14 | 28 | |
| <0.001 | 4989 | 5.1 | -2 | -28 | 45 | PCC |
| | | 4.66 | -4 | -58 | 16 | |
| | | 4.5 | -9 | -16 | 34 | |
| 0.033 | 1024 | 4.79 | -18 | 27 | -21 | OFC |
| | | 3.93 | -26 | 39 | -16 | |
| | | 3.85 | -24 | 46 | -10 | |
| 0.039 | 971 | 4.65 | 39 | -48 | 38 | right IPL |
| | | 4.06 | 42 | -56 | 42 | |
| | | 3.91 | 34 | -52 | 51 | |
| 0.015 | 1279 | 4.44 | 15 | 15 | 14 | caudate |

**Table S4: Brain regions with grey matter volume associated with grey matter volume in the left Inferior Parietal Lobule (IPL).** Initial threshold p<0.001. Abbreviations: Inferior Frontal Gyrus (IFG), Prefrontal Cortex (PFC), Orbitofrontal Cortex (OFC), Posterior Cingulate Cortex (PCC).

| cluster | | peak | Coordinates (MNI) | | | anatomy |
|---|---|---|---|---|---|---|
| p(FWE-corr) | equivk | T | x | y | z | |
| <0.001 | 55887 | 118.74 | -56 | 12 | 15 | seed (left IFG), also containing PFC, PCG |
| | | 6.3 | -6 | 50 | 32 | |
| | | 6.25 | 12 | 69 | 18 | |
| 0.025 | 1084 | 4.86 | 12 | -54 | 16 | PCC |
| | | 4.5 | 21 | -62 | 14 | |
| | | 3.77 | 2 | -62 | 16 | |

**Table S5: Brain regions with grey matter volume associated with grey matter volume in the left Inferior Frontal Gyrus (IFG).** Initial threshold p<0.001. Abbreviations: Inferior Frontal Gyrus (IFG), Prefrontal Cortex (PFC), Precentral Gyrus (PCG), Posterior Cingulate Cortex (PCC).

| cluster | | peak | Coordinates (MNI) | | | anatomy |
|---|---|---|---|---|---|---|
| p(FWE-corr) | equivk | T | x | y | z | |
| <0.001 | 2904 | 70.98 | -44 | 6 | 30 | Seed (left PCG) |

**Table S6: Brain regions with grey matter volume associated with grey matter volume in the left Precentral Gyrus (PCG).** Initial threshold p<0.001.

*Structural covariance matrices in typical-reading children and adults*

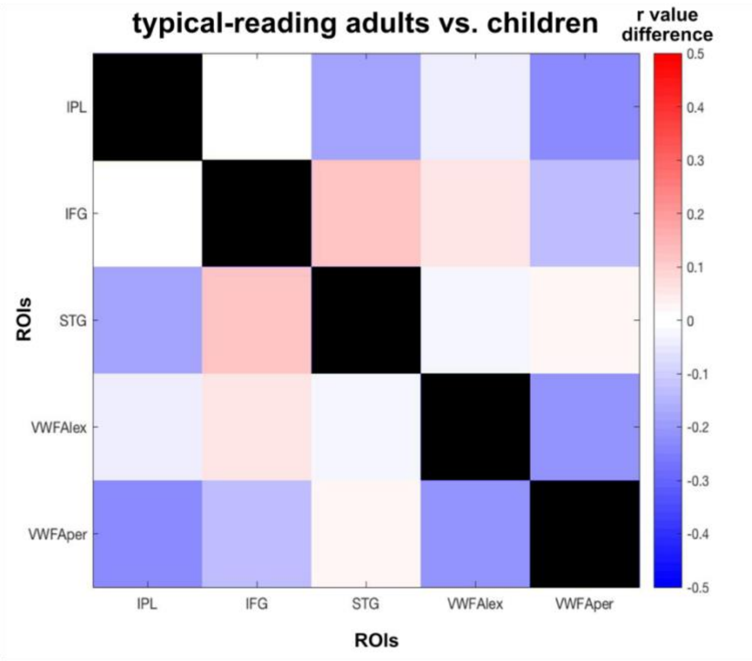

**Figure S1: Difference in structural covariance of reading-related brain regions between typical-reading children and adults.** We did not find a significant difference between typical-reading children and adults for structural covariance of reading-related brain regions. The color of each cell reflects the difference in structural covariance (typical-reading adults minus typical-reading children) of the corresponding column and row ROI.

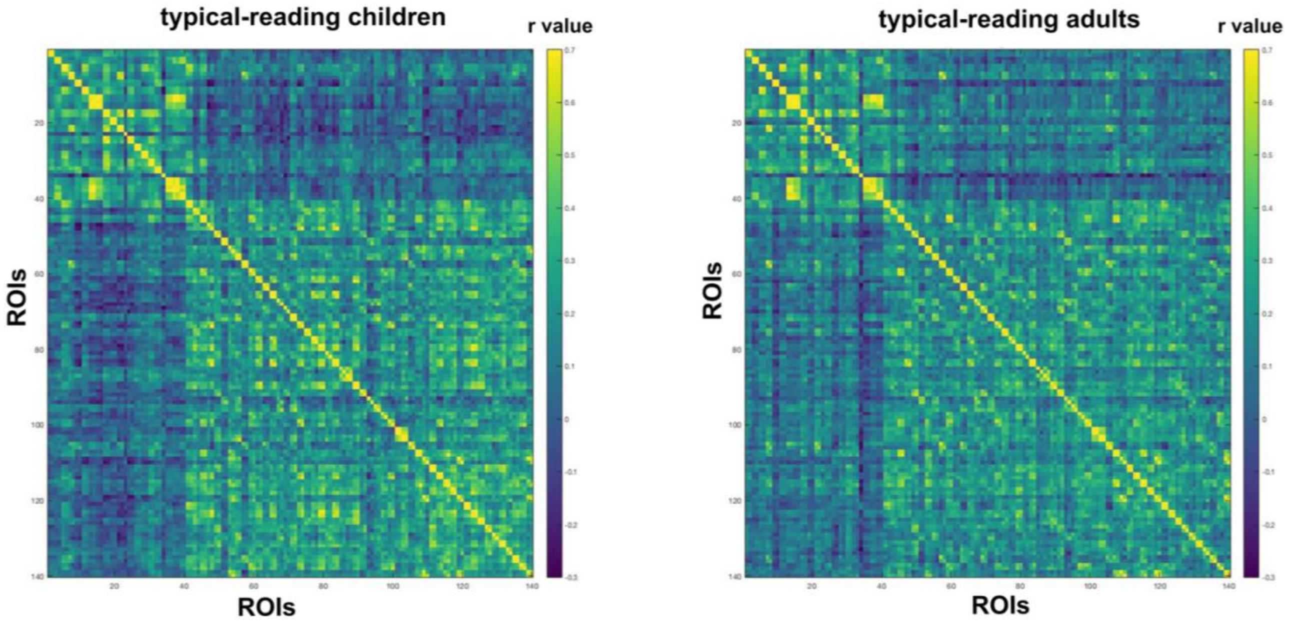


**Figure S2: Structural covariance of 140 ROIs across the whole brain in typical-reading adults and children.** We did not find a significant difference between typical-reading adults and children for structural covariance of the whole brain. The color of each cell reflects the structural covariance of the corresponding column and row ROI.

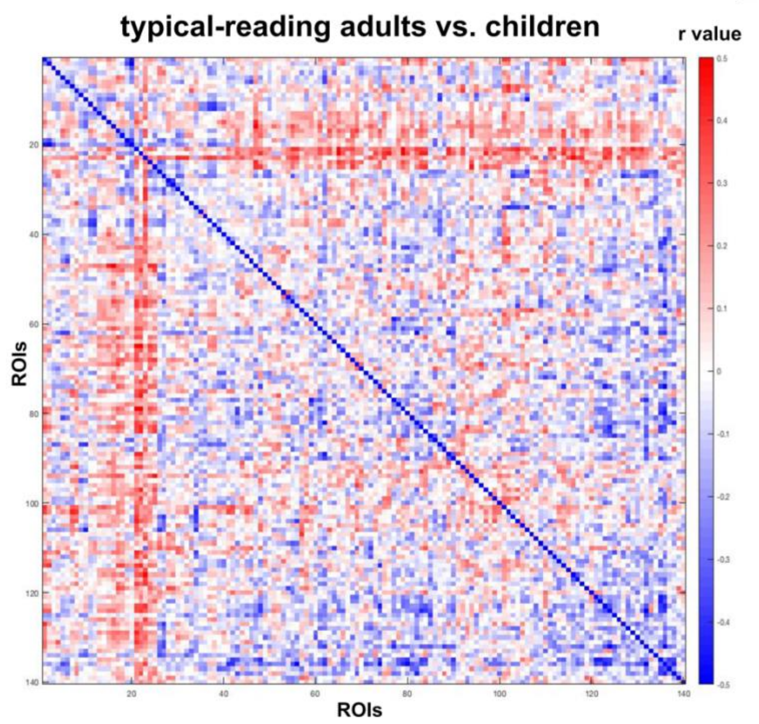


**Figure S3: Difference in structural covariance of 140 ROIs across the whole brain between typical-reading children and adults.** We did not find a significant difference between typical-reading children and adults for structural covariance of 140 ROIs across the whole brain. The color of each cell reflects the difference in structural covariance (typical-reading adults minus typical-reading children) of the corresponding column and row ROI.

*Structural covariance matrices in typical-reading children and children with dyslexia*

For completeness, we also calculated structural covariance matrices for both groups of children using 140 ROIs taken from a whole-brain parcellation. Here, we did not find any significant difference between typical-reading children and children with dyslexia.

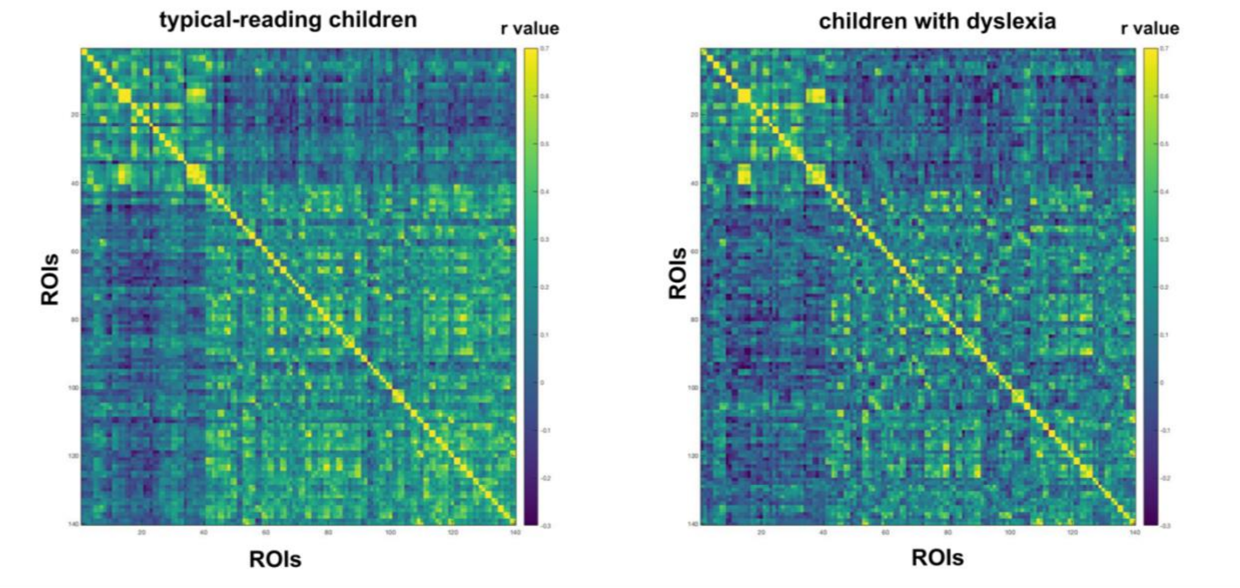


**Figure S4: Structural covariance of 140 ROIs across the whole brain in typical-reading children and children with dyslexia.** We did not find a significant difference between typical-reading children and children with dyslexia for structural covariance of the whole brain. The color of each cell reflects the structural covariance of the corresponding column and row ROI.

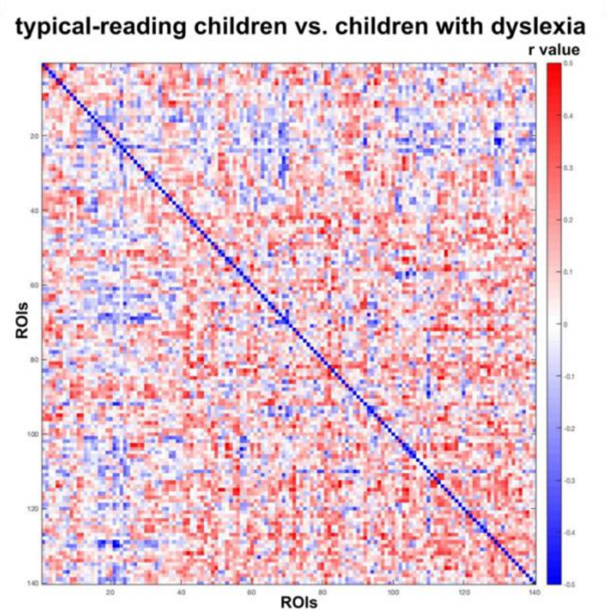


**Figure S5: Difference in structural covariance of 140 ROIs across the whole brain between typical-reading children and children with dyslexia.** We did not find a significant difference between typical-reading children and children with dyslexia for structural covariance of the whole brain. The color of each cell reflects the difference in structural covariance (typical-reading children minus children with dyslexia) of the corresponding column and row ROI.

*Whole-brain seed-based analyses using the lexical and perceptual Visual Word Form Area as seeds in typical-reading children and children with dyslexia*

| cluster | | peak | coordinates (MNI) | | | anatomy |
|---|---|---|---|---|---|---|
| **p(FWE-corr)** | **equivk** | **T** | **x** | **y** | **z** | |
| <0.001 | 3366 | 72.1 | -42 | -58 | -10 | seed (VWFA_lex) |
| | | 6.67 | -44 | -44 | -20 | |
| 0.038 | 906 | 5.47 | 48 | -54 | -12 | right vOTC |
| | | 3.69 | 50 | -33 | -18 | |
| | | 3.34 | 57 | -70 | -6 | |

**Table S7: Brain regions with grey matter volume associated with grey matter volume in the lexical Visual Word Form Area (VWFA_lex) in children with dyslexia.** Initial threshold p<0.001. Abbreviations: ventral occipito-temporal cortex (vOTC).

| cluster | | peak | coordinates (MNI) | | | anatomy |
|---|---|---|---|---|---|---|
| **p(FWE-corr)** | **equivk** | **T** | **x** | **y** | **z** | |
| 0.002 | 1756 | 58.64 | -42 | -72 | -9 | seed (VWFA_per) |
| | | 4.78 | -50 | -62 | -12 | |

**Table S8: Brain regions with grey matter volume associated with grey matter volume in the perceptual Visual Word Form Area (VWFA_per) in children with dyslexia.** Initial threshold p<0.001.

| cluster | | peak | coordinates (MNI) | | | anatomy |
|---|---|---|---|---|---|---|
| **p(FWE-corr)** | **equivk** | **T** | **x** | **y** | **z** | |
| <0.001 | 3514 | 83.14 | -44 | -58 | -10 | seed (VWFA_lex) |
| | | 6.13 | -40 | -45 | -20 | |
| 0.006 | 1603 | 4.9 | 27 | 16 | -15 | insula |
| | | 4.09 | 33 | 20 | -2 | |
| | | 3.84 | 40 | 15 | 4 | |
| 0.058 | 864 | 4.75 | -57 | -34 | 6 | left STG |
| | | 4.27 | -69 | -45 | 9 | |
| | | 3.83 | -45 | -28 | 0 | |

**Table S9: Brain regions with grey matter volume associated with grey matter volume in the lexical Visual Word Form Area (VWFA_lex) in typical-reading children.** Initial threshold p<0.001. Abbreviations: Superior Temporal Gyrus (STG).

| cluster | | peak | cooridnates (MNI) | | | anatomy |
|---|---|---|---|---|---|---|
| **p(FWE-corr)** | **equivk** | **T** | **x** | **y** | **z** | |
| <0.001 | 2549 | 64.58 | -40 | -72 | -8 | left VWFA_per |
| 0.001 | 2136 | 5.01 | 56 | -45 | 42 | right IPL |
| | | 4.41 | 54 | -39 | 52 | |
| | | 4.29 | 56 | -51 | 20 | |

**Table S10: Brain regions with grey matter volume associated with grey matter volume in the perceptual Visual Word Form Area (VWFA_per) in typical-reading children.** Initial threshold p<0.001. Abbreviations: Inferior Parietal Lobule (IPL).
